# A multiethnic GWAS meta-analysis of 585,243 individuals identifies new risk loci associated with cataract and reveals sex-specific effects

**DOI:** 10.1101/2020.09.23.20200428

**Authors:** Hélène Choquet, Ronald B. Melles, Deepti Anand, Jie Yin, Gabriel Cuellar-Partida, Wei Wang, 23andMe Research Team, Thomas J. Hoffmann, K Saidas Nair, Pirro G. Hysi, Salil A. Lachke, Eric Jorgenson

## Abstract

Cataract is the leading cause of blindness among the elderly worldwide and cataract surgery is one of the most common operations performed in the United States^1-3^. The etiology remains largely unclear, and to contribute to its elucidation we conducted a multiethnic genome-wide association meta-analysis of cataract, combining results from the GERA and UK Biobank cohorts, and tested for replication in the research cohort from 23andMe, Inc.. We report 54 genome-wide significant loci, 37 of which were previously unknown. Sex-stratified analyses identified two additional novel loci (*CASP7* and *GSTM2*) specific to women and sex differences in effect sizes and significance of association at five other loci. We show that genes within or near 80% of the cataract-associated loci are significantly expressed and/or enriched-expressed in the mouse lens across various spatiotemporal stages. Further, 32 candidate genes in the associated loci have altered gene expression in 9 different gene perturbation mouse models of lens defects/cataract, suggesting their relevance to lens biology.

---

Cataracts are caused by opacification of the crystalline lens, which leads to progressive loss of vision. They can present as a developmental disorder in younger patients (congenital or pediatric cataracts) but, more commonly, occur as a disease of aging^4,5^, and are a leading cause of visual impairment. Cataract formation and cataract surgery are more common in women^6^. Twin and family aggregation studies strongly support an important role for genetic factors in cataract susceptibility with heritability estimates ranging from 35 to 58%^7-12^. A recent study^13^ investigated the genetic basis of eye disease, reported 20 genetic loci associated with cataract at a genome-wide level of significance in the UK Biobank European sample, although none of these loci was independently replicated. It is also unclear what proportion of clinical variability these loci help explain, as well as to what contribution they have in populations of diverse ethnic background.

Here, we present the largest and most ethnically diverse genetic study of cataract susceptibility conducted to date to our knowledge. Following a stepwise analytical approach, we conducted a genome-wide association analyses, followed by meta-analysis, including 585,243 individuals (67,844 cases and 517,399 cataract-free controls) from two cohorts: the Genetic Epidemiology Research in Adult Health and Aging (GERA)^14^ and the UK Biobank (UKB)^15,16^. We tested the top independently associated SNPs (*P*<5.0×10^−8^) at each locus in 3,234,455 participants (347,209 self-reported cataract cases and 2,887,246 controls) from the 23andMe research cohort. Cohorts summary details are presented in **Supplementary Table 1**. We subsequently fine-mapped these associations^17^ and examined changes in the expression of candidate genes in associated loci in 9 gene perturbation mouse models of lens defects^18,19^. We then undertook conditional, ethnic-, and sex-specific association analyses (**Supplementary Fig. 1**). Finally, we assessed the genetic correlation between cataract and other disorders and complex traits^20^.

We first undertook GWAS analysis of cataract in the GERA and UKB cohorts, stratified by ethnic group, followed by a meta-analysis across all analytical strata. In the multiethnic meta-analysis, we identified 54 loci (*P*<5.0×10^−8^), of which 37 were novel (i.e., not previously reported to be associated with cataract at a genome-wide level of significance) (**Table 1, Fig. 1, and Supplementary Fig. 2**). The effect estimates of 54 lead SNPs were consistent across the 2 studies (**Table 1 and Supplementary Fig. 3**). In 23andMe research cohort, 45 out of 51 lead SNPs available (88.2%) replicated with a consistent direction of effect at a Bonferroni corrected significance threshold of 9.8×10^−4^ (*P*-value=0.05/51) and additional 2 SNPs were nominally significant (*P*<0.05) (**Table 1 and Supplementary Fig. 4**).

**Table 1.**
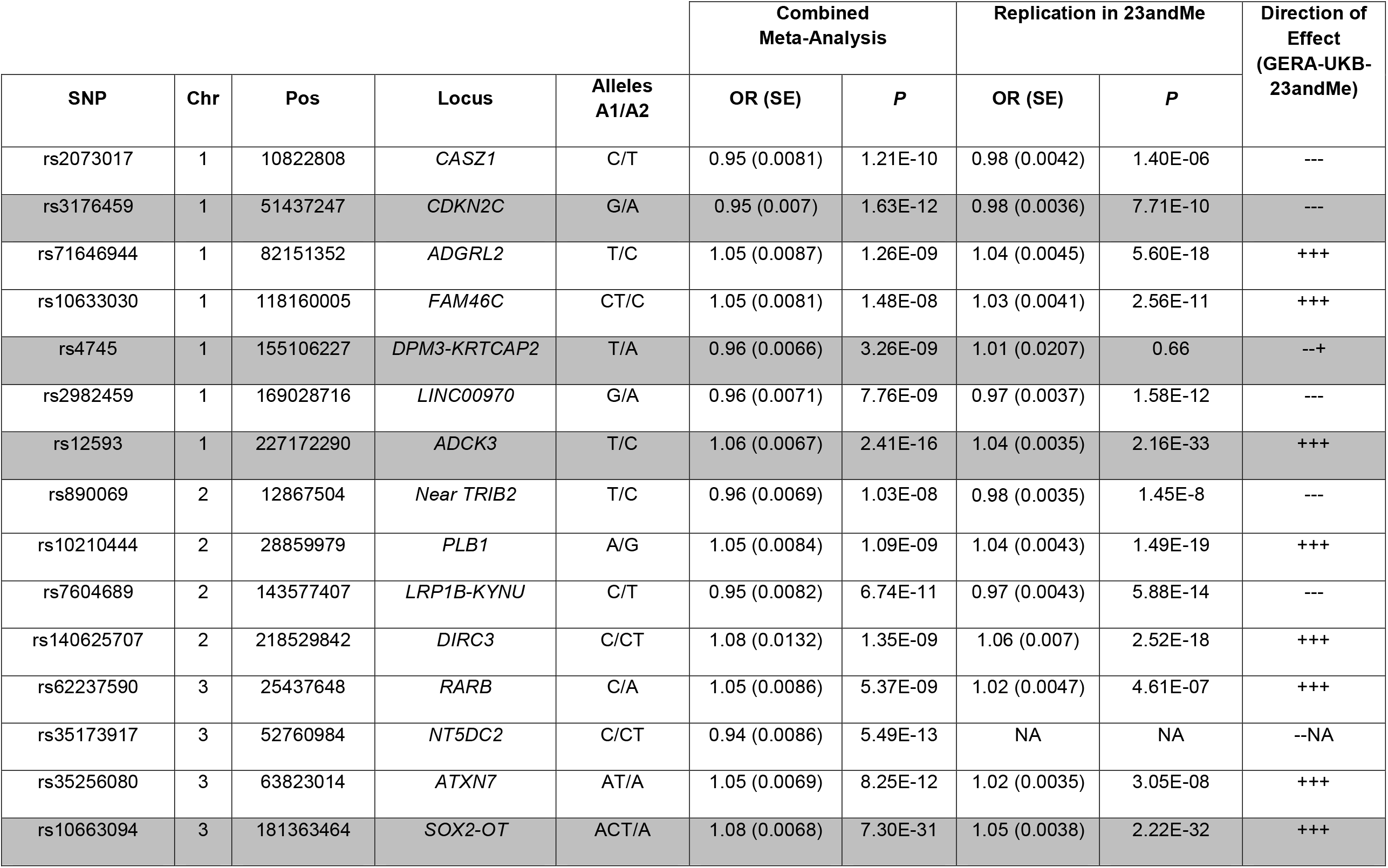
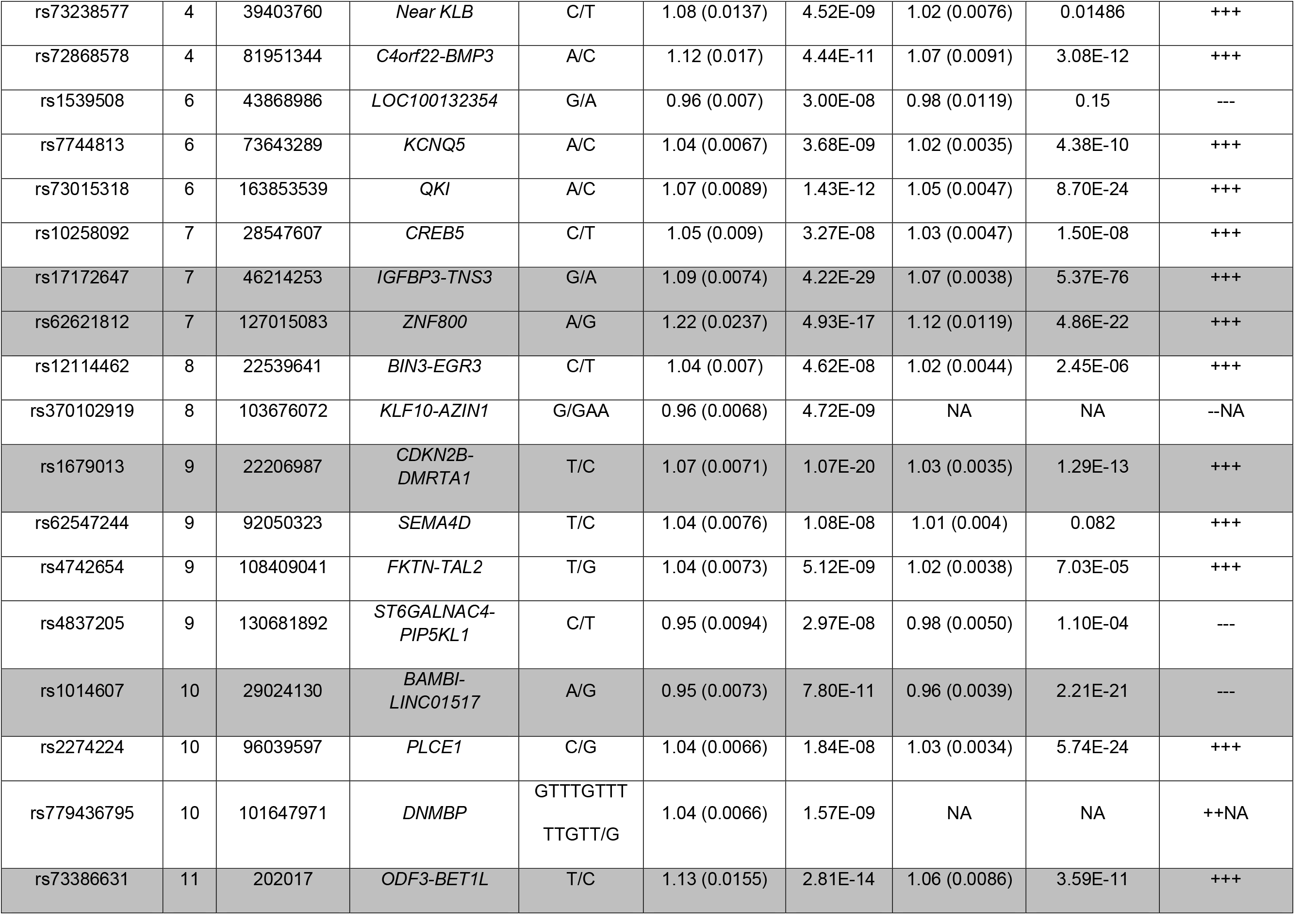
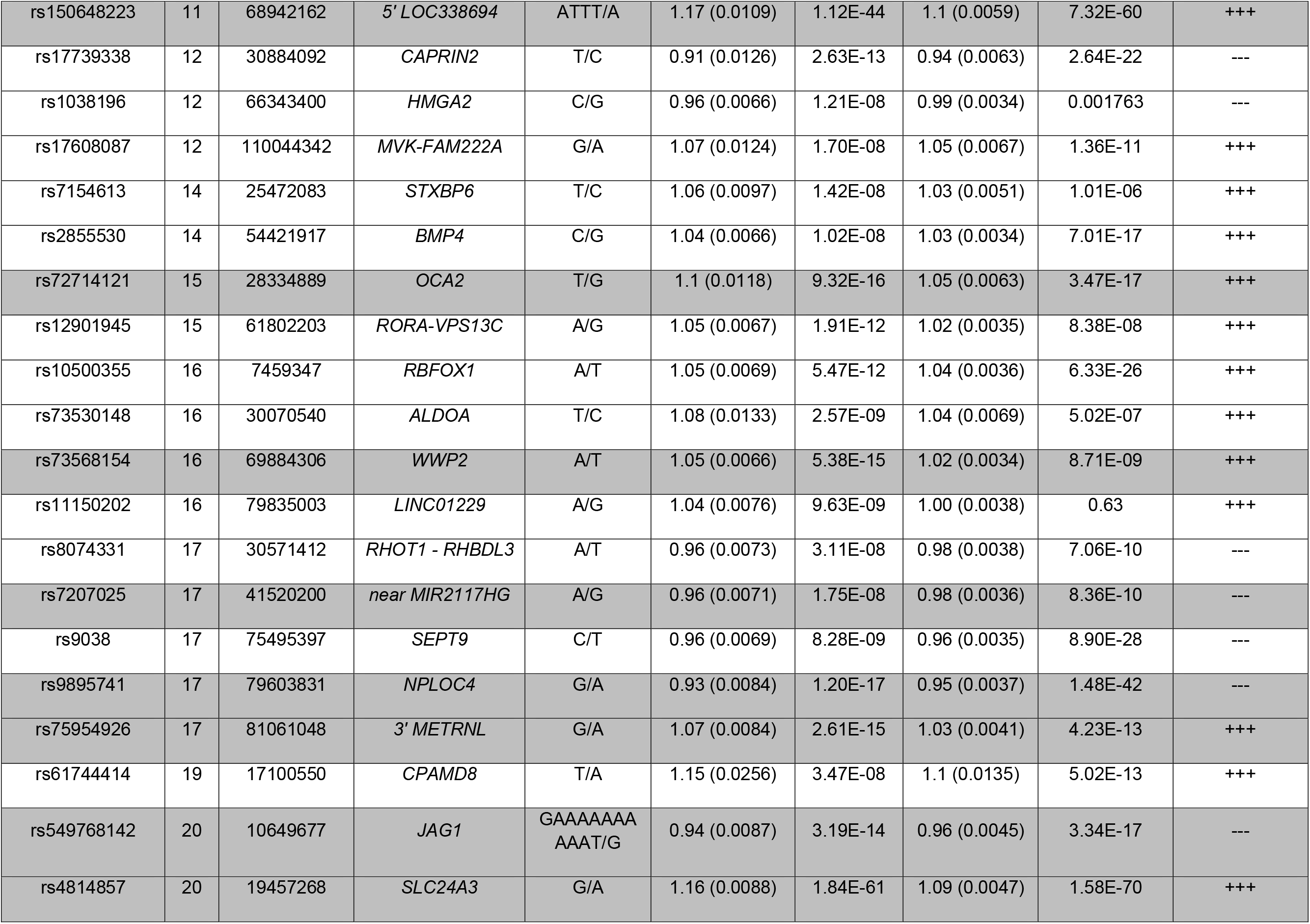
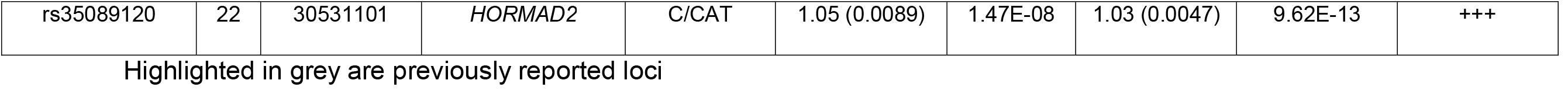
Cataract loci identified in the combined (GERA+UKB) GWAS multiethnic meta-analysis and replication in 23andMe research cohort

**Figure 1.**
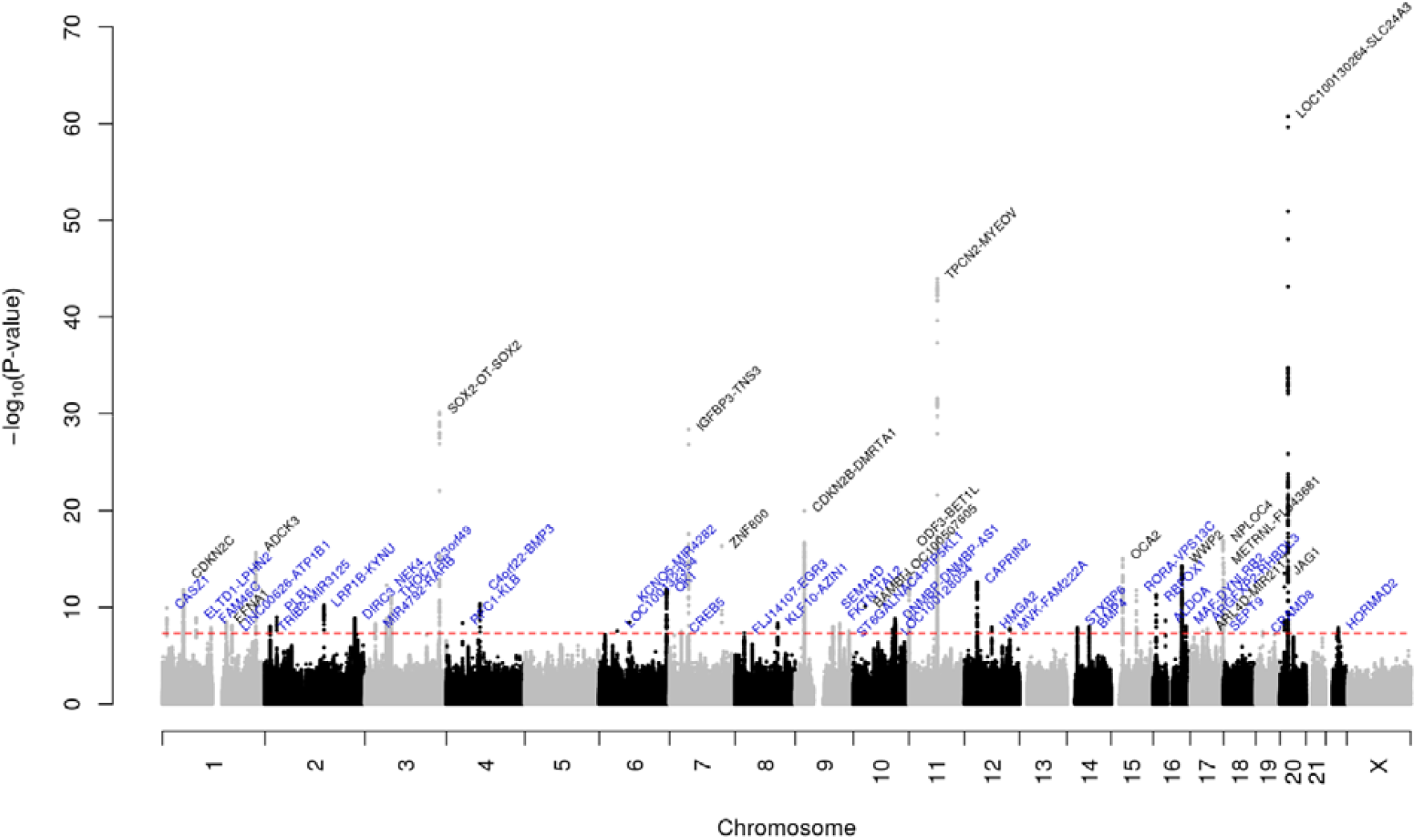
Manhattan plot of the multiethnic combined (GERA+UKB) GWAS meta-analysis of cataract. Locus names in blue are for the novel loci and the ones in dark are for the previously reported ones.

To determine whether there were additional signals in individual ancestry groups that did not reach genome-wide significance in the meta-analysis, we conducted ethnic-specific meta-analyses of each ancestry group. We identified three additional novel loci in the European ancestry (GERA non-Hispanic whites + UKB Europeans) meta-analysis: *EPHA4, CD83-JARID2*, and near *EXOC3L2* (**Supplementary Fig. 5.a. and Supplementary Table 2**). Regional association plots of the association signals are presented in **Supplementary Fig. 6**. To identify independent signals within the 44 genomic regions identified in the European-specific meta-analysis (**Supplementary Table 3**), we performed a multi-SNP-based conditional & joint association analysis (COJO)^21^, which revealed 5 additional independent SNPs within 4 of the identified genomic regions, including at known loci (*CDKN2B, RIC8A*, and *LOC338694*) and at newly identified *DNMBP* locus (**Supplementary Table 4**). Neither the meta-analysis of East Asian groups nor the meta-analysis combining the GERA African American and UKB African British groups result in the identification of additional novel genome-wide significant findings (**Supplementary Fig. 5.b. and 5.c**.).

Next, we conducted genetic association analyses for interaction between genetic factors and sex, in sex-specific meta-analyses combining data from GERA and UKB. We identified two additional novel loci, *CASP7* and *GSTM2*, in the women-specific meta-analysis (GERA+UKB) (**Fig. 2 and Supplementary Table 5**). *CASP7* rs12777332 and *GSTM2* rs3819350 were significantly associated with cataract in women (*CASP7* rs12777332: OR=1.06, *P*=3.41×10^−8^; *GSTM2* rs3819350: OR=1.06, *P*=2.10×10^−8^) but not in men (*CASP7* rs12777332: OR=1.01, *P*=0.25; *GSTM2* rs3819350: OR=1.01, *P*=0.25) (**Supplementary Fig. 7**). Further, among the loci identified in the multiethnic meta-analysis (GERA+UKB), we observed significant differences in the effect sizes and significance of association at five loci: one locus, *DNMBP-CPN1*, was strongly associated with cataract in women but not in men (*DNMBP-CPN1* rs1986500, OR=0.94, *P*=5.04×10^−11^ in women, and OR=1.01, *P*=0.40 in men; *Z*=-5.03, *P*=2.44×10^−7^) (**Supplementary Fig. 8 and Supplementary Table 5**); and four loci, *QKI, SEMA4D, RBFOX1*, and *JAG1*, were strongly associated in men than women (*QKI* 6:163840336, OR=0.94, *P*=1.23×10^−10^ in men, and OR=0.99, *P*=0.21 in women; *Z*=-3.95, *P*=3.89×10^−5^; *SEMA4D* rs62547232, OR=1.15, *P*=1.83×10^−9^ in men, and OR=0.98, *P*=0.33 in women; *Z*=5.03, *P*=2.43×10^−7^; *RBFOX1* rs7184522, OR=1.07, *P*=9.10×10^−12^ in men, and OR=1.03, *P*=0.0020 in women; *Z*=2.98, *P*=1.43×10^−3^; *JAG1* rs3790163, OR=0.92, *P*=3.14×10^−12^in men, and OR=0.96, *P*=9.63×10^−4^ in women; *Z*=-2.95, *P*=1.59×10^−3^) (**Fig. 2 and Supplementary Table 6 and Supplementary Fig. 8**). Regional association plots illustrate the sex-specific association signals (**Supplementary Fig. 7**).

**Figure 2.**
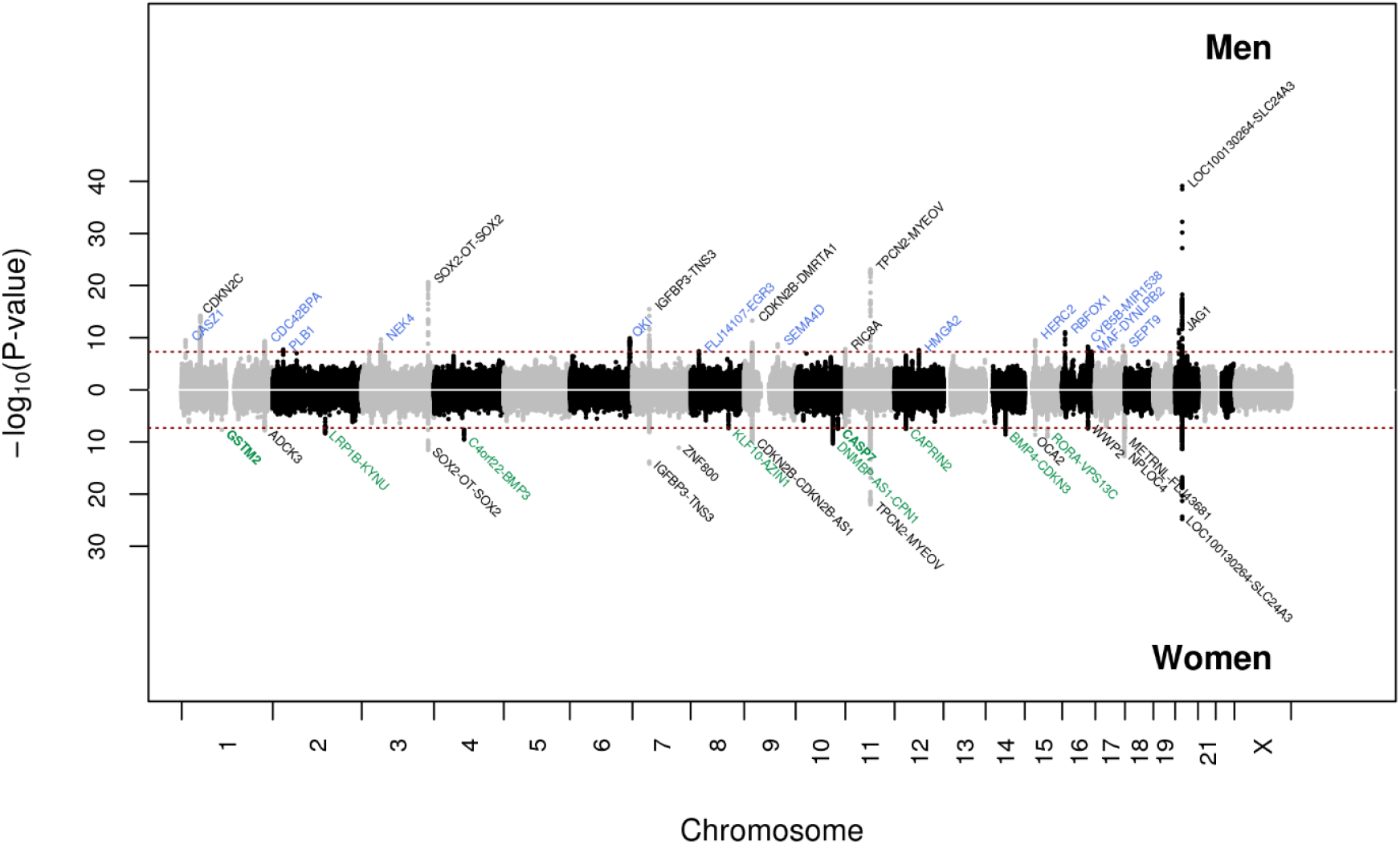
Chicago plot of the sex-stratified multiethnic GWAS meta-analyses of cataract, combining GERA and UKB in men (upper panel) and women (lower panel) Locus names in black are for those previously reported. Locus names in bold (*CASP7* and *GSTM2*) are for the additional novel loci specific to women (compared to the multiethnic meta-analysis (GERA+UKB)). Novel loci significantly associated (*P* <5 × 10^−8^) with cataract in women are highlighted in green, and those significantly associated with cataract in men are highlighted in blue.

We adopted a Bayesian approach (CAVIARBF)^17^ to compute variants likelihood to explain the observed association at each locus and derived the smallest set of variants that has a 95% probability to include the causal origin of the signals (95% credible set). Nine sets included a single variant (**Supplementary Table 7**) such as rs62621812 (*ZNF800*), rs1014607 (*BAMBI-LOC100507605*), rs1428885924 (*NEK4*), rs1679013 (*CDKN2B-DMRTA1*), rs1539508 (*LOC100132354*), rs73238577 (*RFC1-KLB*), rs17172647 (*IGFBP3-TNS3*), rs73530148 (*ALDOA*), and rs549768142 (*JAG1*), suggesting that those single variants may be the causal origin of the associations observed in their respective loci.

A gene-based analysis, using the VEGAS2 integrative tool^22^ on 22,673 genes, found significant associations with cataract for 8 genes within 4 loci identified in the multiethnic combined (GERA+UKB) meta-analysis, including *EFNA1* and *KRTCAP2* (chr1q22), *CDKN2B* and *CDKN2B-AS1* (chr9p21.3), *MRPL21* and *LOC338694* (chr11q13.3), *HERC2* (chr15q13.2), and *BLVRA* gene (chr7p13) (**Supplementary Table 8**).

We next examined the expression of genes within identified loci potentially associated with cataract in lens tissue using the web-resource tool iSyTE (integrated <u>Sy</u>stems <u>T</u>ool for <u>E</u>ye gene discovery)^18,19^. iSyTE contains genome-wide expression data, based on microarray or RNA-seq analysis, on the mouse lens at different embryonic and postnatal stages^18,23^. In addition to expression, iSyTE also contains information of “lens-enriched expression” which has proved to be an excellent predictor of cataract-linked genes in humans and animal models^24-27^. The iSyTE-based lens microarray data on Affymetrix and/or Illumina platforms showed that orthologs of 47 candidates were significantly expressed in the mouse lens (>100 expression units, *P*<0.05) in one or more embryonic/postnatal stages (**Fig. 3**). Over 60% of the expressed genes were found to have high lens-enriched expression (>1.5 fold-change over whole embryonic body (WB) reference dataset, *P*<0.05), suggesting their likely relevance to lens development, homeostasis and pathology (**Supplementary Fig. 9**). This was further supported by iSyTE RNA-seq data that also showed lens-expression of 46 candidates (≥2.0 CPM, counts per million, *P*<0.05), 31 out of which (∼68%) exhibited high lens-enriched expression in one or more embryonic/postnatal stages (>1.5 fold-change over WB, *P*<0.05) (**Fig. 3 and Supplementary Fig. 9**). Together, this analysis offered strong support for lens expression for total 52 different genes, with at least one candidate gene for 43 of the 54 loci, thus accounting for nearly 80% of the identified loci. Additionally, iSyTE also informs on lens gene expression changes in specific gene perturbation mouse models of lens defects/cataract. This analysis showed that 38 candidate genes had significant differences in gene expression (*P*<0.05) in one or more of the 9 different gene perturbation mouse models of lens defects/cataract (**Supplementary Fig. 10 and Supplementary Table 9**). Together, iSyTE analysis offers independent experimental evidence that support the direct relevance of these candidate genes to lens biology and cataract.

**Figure 3.**
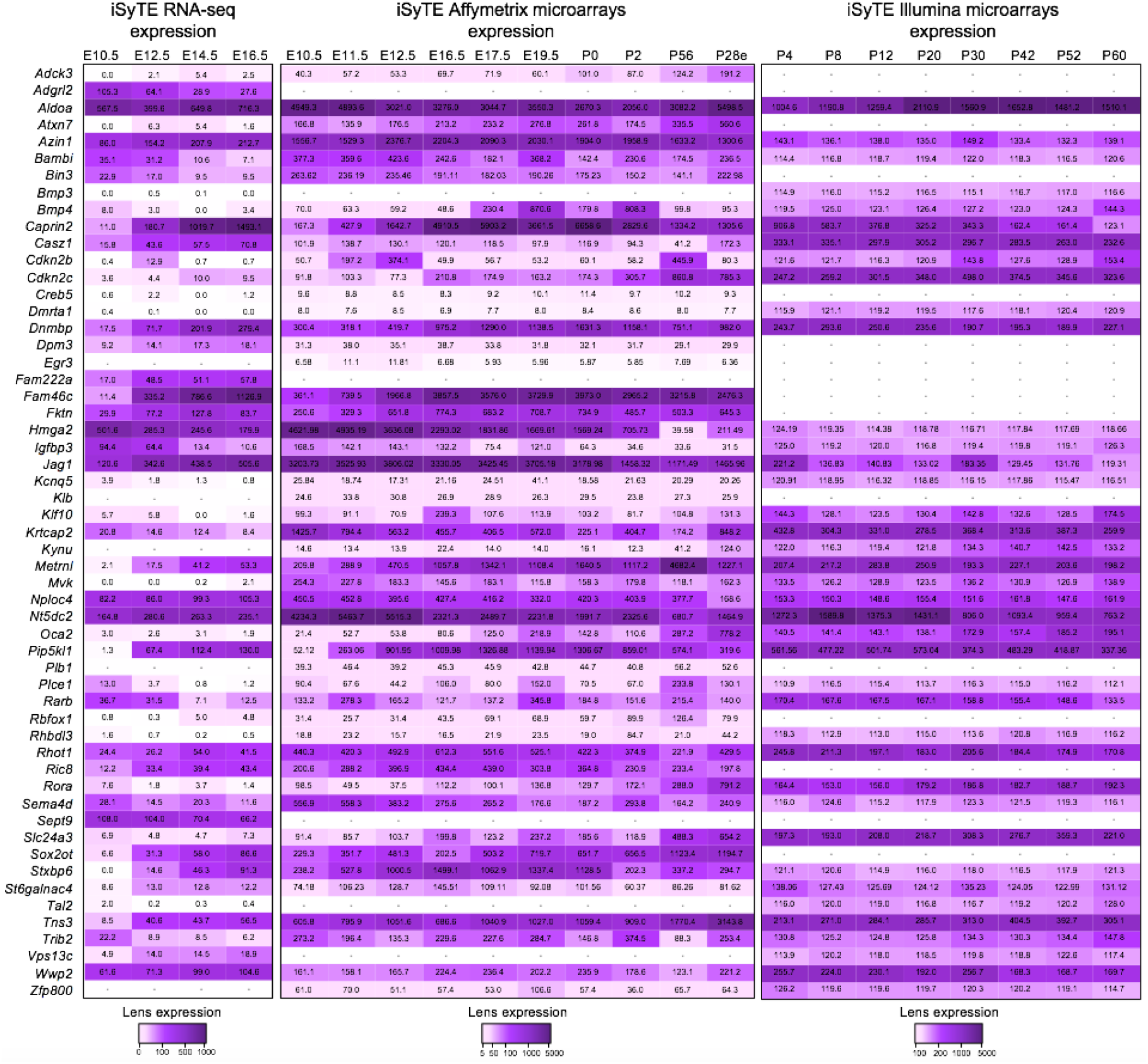
Expression of candidate genes in mouse lens. Mouse orthologs of the human candidate genes in the 54 loci were examined for their lens expression in the iSyTE database. Analysis of whole lens tissue data on various platforms, microarrays (Affymetrix, Illumina) and RNA-seq indicates expression of 55 genes at different stages indicated by embryonic (E) and postnatal (P) days and ranged from early lens development (*i*.*e*., E10.5) through adulthood (*i*.*e*. P60). Note: P28 in Affymetrix represents expression data on isolated lens epithelium. The range of expression on each platform is indicated by a specific heat-map. The numbers within individual tiles indicate the level of expression in fluorescence intensity (for microarrays) and in counts per million (CPM) (for RNA-seq).

We also conducted a pathway analysis using VEGAS software^22^ to assess enrichment in 9,732 pathways or gene-sets derived from the Biosystem’s database. We found that the notochord development was the only gene-set significantly enriched in our results, after correcting for multiple testing (*P*<5.14×10^−6^) (**Supplementary Table 10**). In addition, we identified 781 pathways/gene-sets that were nominally enriched (*P*<0.05), with the most significant of which were ‘circadian clock’ (*P*=2.07×10^−5^), followed by ‘lens morphogenesis in camera-type eye’ (*P*=2.14×10^−5^), and ‘notochord morphogenesis” (*P*=2.81×10^−5^). Our findings are consistent with early work, demonstrating that mice deficient in circadian clock proteins, such as *BMAL1* and *CLOCK*, display age-related cataract^28,29^.

To estimate the pairwise genetic correlations (r_g_) between cataract and more than 700 diseases/traits from different publicly available resources/consortia, we compared our GWAS results with summary statistics for other traits by performing an LD score regression using the LD Hub web interface^20^. Genetic correlations were considered significant after Bonferroni adjustment for multiple testing (*P*<6.48×10^−5^ which corresponds to 0.05/772 phenotypes tested). We found significant genetic correlations between cataract and 39 traits, including three of them directly related to eye traits: ‘wears glasses or contact lenses’ (r_g_=0.30, *P*=2.56×10^−7^), ‘self-reported: glaucoma’ (r_g_=0.30, *P*=4.57×10^−6^), and ‘reason for glasses/contact lenses: myopia’ (r_g_=0.25, *P*=1.10×10^−5^) (**Supplementary Table 11**).

A phenome-wide association study (PheWAS) analysis of 43 cataract-associated SNPs, available in the GeneATLAS was run across 776 traits measured and previously analyzed in the UKB^30^. Twenty-three of the most significantly associated cataract-associated variants were significantly associated (*P*<5.0×10^−8^) with other traits (**Fig. 4)**. Most were associated with disorders of the lens, with the strongest association observed for the intronic variant rs4814857 at *SLC24A3* (*P*=2.48×10^−39^) (**Supplementary Table 12**). *SLC24A3* encodes the carrier family 24 member 3 and has been thought to be involved in retinal diseases^31^. Variants at *PLCE1* and *HMGA2* were significantly associated with anthropometric traits, such as impedance of whole body, impedance of arm and leg. The *PLCE1* gene encodes the phospholipase C epsilon 1 and common variation at this locus has been shown to be associated with retinal detachment^13^ (**Supplementary Table 13**), and *HMGA2* loss of function variants were linked with bilateral cataracts in a fetus presenting with growth delay^32^. Our PheWAS analysis also highlighted that variants at *OCA2* and *NPLOC4* were significantly associated with pigmentation phenotypes. The *OCA2* gene encodes the melanosomal transmembrane protein, whose variants determine iris color and have been linked to corneal and refractive astigmatism, syndromic forms of myopia, refractive error, and type 2 oculocutaneous albinism^33-37^ (**Supplementary Table 13**). *NPLOC4* encodes the homolog, ubiquitin recognition factor and has been previously associated with macular thickness and the risk of strabismus and corneal and refractive astigmatism^34,38,39^.

**Figure 4.**
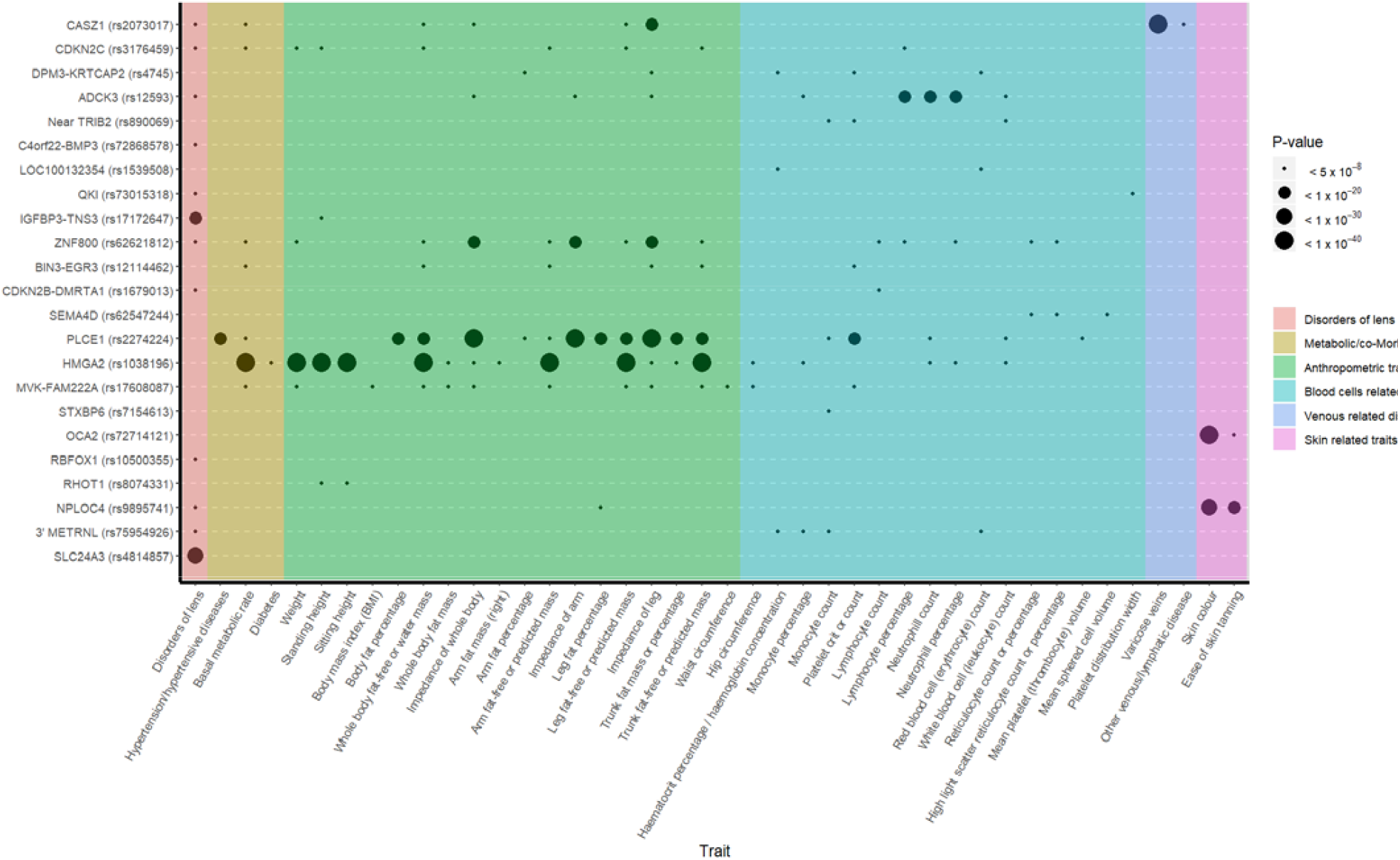
Phenome-wide association matrix of cataract top variants. PheWAS was carried out for the 54 lead SNPs in our loci of interest identified in the combined (GERA+UKB) multiethnic analysis. SNPs were queried against 776 traits ascertained for UKB participants and reported in the Roslin Gene Atlas^30^, including disorders of the lens, anthropometric traits, hematologic laboratory values, ICD-10 clinical diagnoses and self-reported conditions. Among the 54 lead SNPs, 43 were available in Gene Atlas database. We reported SNPs showing genome-wide significant association with at least one trait (in addition to cataract).

Our study should be interpreted within the context of its limitations. First, the cataract phenotypes were assessed differently across the 3 study cohorts. While our cataract phenotype in GERA was based on electronic health records (EHRs) data and International Classification of Disease, Ninth (ICD9) or Tenth Revision (ICD10) diagnosis codes, most of the cataract cases in UKB, and all of the cataract cases in 23andMe research cohort (our replication sample) were based on self-reported data. This may result in phenotype misclassification, however, our meta-analysis combining GERA and UKB showed consistency of the SNPs effect estimates between cohorts, and the identified associations were well validated in the 23andMe research cohort. Second, subtypes of cataract were not available in the 3 study cohorts, which may result in underestimates of the effects of individual SNPs due to phenotype misclassification. Future studies will determine whether the identified loci contribute to different cataract subtypes (i.e. nuclear, cortical, or subcapsular) and the extent to which these loci display shared effects across subtypes.

In conclusion, we report the results of a large GWAS that identified 47 novel loci (37 from the multiethnic-meta-analysis + 3 European-specific meta-analysis + 5 conditional analysis + 2 from the female-specific meta-analysis) for the development of cataract and that likely contribute to the pathophysiology of this common vision disorder. Several genes within these cataract-associated loci, including *RARB, KLF10, DNMBP, HMGA2, MVK, BMP4, CPAMD8*, and *JAG1*, represent potential candidates for the development of drug targets as previous work supports the relevance of these candidates to cataracts^32,40-46^ (**Supplementary Table 13**). We also report three loci that show women-specific effects on cataract susceptibility and 4 others that showed significant differences in effects between women and men. These loci provide a biological foundation for understanding the etiology of sex-differences in cataract susceptibility, and, may suggest potential targets for the development of non-surgical treatment of cataracts.

## Data Availability

The GERA genotype data are available upon application to the KP Research Bank (https://researchbank.kaiserpermanente.org/). The combined (GERA+UKB) meta-analysis GWAS summary statistics are available from the NHGRI-EBI GWAS Catalog (https://www.ebi.ac.uk/gwas/downloads/summary-statistics). The variant-level data for the 23andMe replication dataset are fully disclosed in the manuscript. Individual-level data are not publicly available due participant confidentiality, and in accordance with the IRB-approved protocol under which the study was conducted.

## Acknowledgements

We are grateful to the Kaiser Permanente Northern California members who have generously agreed to participate in the Kaiser Permanente Research Program on Genes, Environment, and Health. Support for participant enrollment, survey completion, and biospecimen collection for the RPGEH was provided by the Robert Wood Johnson Foundation, the Wayne and Gladys Valley Foundation, the Ellison Medical Foundation, and Kaiser Permanente Community Benefit Programs. Genotyping of the GERA cohort was funded by a grant from the National Institute on Aging, National Institute of Mental Health, and National Institute of Health Common Fund (RC2 AG036607). H.C. and E.J. were supported by the National Eye Institute (NEI) grant R01 EY027004, the National Institute of Diabetes and Digestive and Kidney Diseases (NIDDK) R01 DK116738 and by the National Cancer Institute (NCI) R01CA2416323. This work was also made possible in part by NIH-NEI EY002162—Core Grant for Vision Research, by the Research to Prevent Blindness Unrestricted Grant (UCSF, Ophthalmology). K.S.N. receives support from NEI grant EY022891, BrightFocus Foundation (G2019360), Marin Community Foundation-Kathlyn McPherson Masneri and Arno P. Masneri Fund, and That Man May See Inc. T.J.H. was supported by National Institutes of Aging (NIA) grant R21 AG046616. S.A.L. was supported by National Institutes of Health / National Eye Institute grants R01 EY021505 and EY029770 and D.A. was supported by a Knights Templar Pediatric Ophthalmology Career Starter Grant Award. We would like to thank the research participants and employees of 23andMe for making this work possible. The following members of the 23andMe Research Team contributed to this study: Michelle Agee, Stella Aslibekyan, Adam Auton, Elizabeth Babalola, Robert K. Bell, Jessica Bielenberg, Katarzyna Bryc, Emily Bullis, Briana Cameron, Daniella Coker, Gabriel Cuellar Partida, Devika Dhamija, Sayantan Das, Sarah L. Elson, Teresa Filshtein, Kipper Fletez-Brant, Pierre Fontanillas, Will Freyman, Pooja M. Gandhi, Karl Heilbron, Barry Hicks, David A. Hinds, Karen E. Huber, Ethan M. Jewett, Yunxuan Jiang, Aaron Kleinman, Katelyn Kukar, Keng-Han Lin, Maya Lowe, Marie K. Luff, Jennifer C. McCreight, Matthew H. McIntyre, Kimberly F. McManus, Steven J. Micheletti, Meghan E. Moreno, Joanna L. Mountain, Sahar V. Mozaffari, Priyanka Nandakumar, Elizabeth S. Noblin, Jared O’Connell, Aaron A. Petrakovitz, G. David Poznik, Anjali J. Shastri, Janie F. Shelton, Jingchunzi Shi, Suyash Shringarpure, Chao Tian, Vinh Tran, Joyce Y. Tung, Xin Wang, Wei Wang, Catherine H. Weldon, Peter Wilton.

## Author contributions

H.C., S.A.L., and E.J. contributed to study conception and design. T.J.H., and E.J. were involved in the genotyping and quality control of the GERA samples. T.J.H. performed the imputation analyses in the GERA cohort. R.B.M. extracted phenotype data for the GERA subjects based on EHRs. J.Y. performed statistical analyses and in silico analyses. D.A., and S.A.L. carried out the gene expression analyses in lens tissue using iSyTE. W.W. and G.C.P. oversaw the replication analyses in the 23andMe research cohort. H.C., S.A.L., and E.J. interpreted the results of analyses and wrote the manuscript with help from R.B.M., K.S.N., P.G.H.

